# Hospital Care Trajectories of Older Adults with Cancer and the Associated Clinical Profiles: linking a French Prospective Cohort Study and a Clinical Data Warehouse

**DOI:** 10.1101/2024.02.02.24302125

**Authors:** Charline Jean, Elena Paillaud, Pascaline Boudou-Rouquette, Claudia Martinez-Tapia, Frédéric Pamoukdjian, Meoïn Hagège, Stéphane Bréant, Claire Hassen-Khodja, Pierre-André Natella, Tristan Cudennec, Marie Laurent, Philippe Caillet, Etienne Audureau, Florence Canouï-Poitrine

## Abstract

**Background:** The longitudinal hospital care experiences of older adults with cancer, from the treatment decision-making process until their end of life, remain unexplored. We examined the hospital care trajectories of these patients and identified associated clinical determinants.

**Methods:** We linked the ELCAPA multicenter cohort study (patients aged ≥70 with a solid tumor and having been referred for a geriatric assessment between 2012 and 2019) and the Greater Paris University Hospitals’ clinical data warehouse. Individual care trajectories, defined as series of consultations, hospital admissions (in day, acute or rehabilitation units) and emergency room visits, were clustered using multichannel sequence analysis. Cluster membership determinants were identified among socio-demographic, oncological and geriatric parameters by logistic regression analysis.

**Results:** 707 patients (median age: 82; metastatic cancer: 45.2%; 10,998 care episodes) were included. Four trajectory clusters were identified: cluster A (n=149, 21.1%) with in-hospital surgical trajectories, cluster B (n=198, 28.0%) with outpatient care trajectories with chemotherapy and/or radiotherapy, cluster C (n=302, 42.7%) without any hospital cancer treatments, and cluster D (n=58, 8.2%) with mostly chemotherapy and high hospital care consumption. Cluster belonging determinants included metastatic status and cancer site (for cluster A), cognition, mobility and mood status (unimpaired parameters for cluster B and impaired for cluster C), and younger age (for cluster D).

**Conclusion:** While highlighting varied hospital care experiences among older patients with cancer, we found that age remains an independent determinant of chemotherapy-dominant care trajectories.

## Introduction

Patients aged 70 or over account for more than a third of the overall cancer population (1). Despite continuous efforts in the last decades to increase the inclusion rate of older cancer patients in clinical trials, they are still underrepresented(2, 3). Included patients typically have fewer geriatric impairments and comorbidities than the average patients of their age group(4). As a result, making treatment decision for older patients with cancer in clinical setting is complex, and the delivery of care is often empirical.

Analyzing the real-world experiences of older patients with cancer could help bridging this evidence gap(5), but most of the data published on care trajectories in geriatric oncology jumps from the treatment decision-making process to the last month of life(6-9). There is yet a scarcity of data on the patient’s journey from the period before the first geriatric assessment prior to a decision on cancer treatment up to 12 months after inclusion. Such information is important to identify the geriatric and oncological characteristics associated with care trajectories, to detect potentially underserved older patients’ populations and guide more tailored healthcare programs.

This paucity of data arguably pertains to the difficulty to access datasets that combine both detailed information on the care trajectories and refined description of the geriatric and oncological characteristics of the patients. In that regard, claims data based on the retrospective data collection from electronic health records (EHRs) are particularly useful guides to care trajectories but provide little information on the patient’s clinical profile(10-12). Conversely, clinical cohorts enable the prospective collection of detailed data on the patients’ clinical phenotypes but usually lack information on care consumption. We hypothesized that the combination of claims and clinical databases would provide a unique opportunity to accurately depict the care trajectories of older adults with cancer in the light of their clinical profiles. Hence, the objectives of the present study of older patients with cancer were to describe the real-life hospital care trajectories of older cancer patients and identify their clinical determinants.

## Methods

### Data sources, study design and setting

We conducted a retrospective cohort study by linking the Elderly Cancer Patients (ELCAPA) cohort study’s database to the clinical data warehouse (CDW) curated by the Greater Paris University Hospitals Group (Assistance Publique – Hôpitaux de Paris, AP-HP, Paris, France).

The AP-HP’s CDW gathers data from EHRs of 39 Paris area public-sector hospitals aggregated into six hospital groups. These EHRs include claims data (diagnoses from the International Classification of Diseases, 10th revision (ICD-10), medical procedures in disease-related groups (DRGs), clinical reports, and information on the units and wards attended during in- and outpatient care (the type of unit/ward, the admission and discharge dates, and admission and discharge modes). In order to define our study’s start date, we estimated the CDW’s data completeness dates for each of the six hospital groups (see Supplementary Materials – SM 1 for details). The AP-HP’s CDW was registered with the French National Data Protection Commission (CNIL, Paris, France; reference: 1980120, January 2017).

The ELCAPA prospective, multicenter cohort study was initiated in 2007. It includes cancer patients aged 70 or over having been referred for a geriatric assessment (GA) to a geriatric oncology clinic in the Paris area (19 centers, including 13 from the AP-HP) prior to a decision on cancer treatment. Data on oncological features, comorbidities, geriatric domains, and sociodemographic parameters are collected. The study has been registered at ClinicalTrials.gov (identifier: NCT02884375) and has been described in detail elsewhere (13). The study protocol was approved by an institutional review board (*CPP Ile-de-France I*, Paris, France; reference: May 2019-MS121). All patients provided their verbal, informed consent before inclusion.

The present study was approved by the AP-HP’s CDW institutional review board (IRB 00011591, Paris, France; reference: CSE 21-49). The research was reported on in compliance with the Reporting of Studies Conducted using Observational Routinely-collected Data (RECORD) guidelines (checklist given in SM 2).

### Study population

We linked the ELCAPA clinical database to the AP-HP’s CDW by directly matching patients via the AP-HP’s unique patient identifier, which is recorded manually upon at enrollment in the ELCAPA study. To ensure that the linkage was accurate, each patient’s initials, sex, and date of birth were cross-checked.

#### Study sample

The study sample included all correctly matched patients with a solid tumor enrolled in an ELCAPA AP-HP center between the hospital group’s data completeness date and March 2019. Patients with hematologic cancer usually receive specific treatments, follow dedicated pathways differing from solid tumors, and were consequently excluded. We limited our analysis to the period before the epidemic of coronavirus disease 2019 because it led to significant disruptions of hospital care trajectories - especially among older patients (14). Sensitivity analyses were performed by limiting the study sample to patients newly diagnosed with cancer (a feature recorded in ELCAPA) and therefore excluding those who were already being treated on the inclusion date.

#### Follow-up

Patients were followed up from two months before inclusion in the ELCAPA study (during which the GA occurred, giving the index date) until 12 months after inclusion. Cancer diagnoses and the initiation of treatment might have occurred for some patients a few weeks before their inclusion in the ELCAPA study, hence the two months look back. Patients without at least one hospital visit documented in the CDW during this period were excluded. Patients’ vital status and early death, i.e. death within 100 days from the index date (15), were recorded.

### Data collection

#### Care episodes

All care episodes (broadly defined as meaningful units of analysis for assessing the full range of services provided in treating a particular health problem (16)) were extracted from the CDW and categorized as consultations, day hospital admissions, acute ward admissions, rehabilitation unit admissions, or emergency room visits. Hospital admissions were also characterized by their reason for admission (cancer-related or not), using a tagging algorithm developed by the French National Cancer Institute. This algorithm demonstrated positive and negative predictive values of 90.5% and 97.5%, respectively, as reported in their validation study (17). This led to the categorization of care episodes into seven dimensions, which were further divided into two to six subdimensions (Table 1).

**Table 1.**
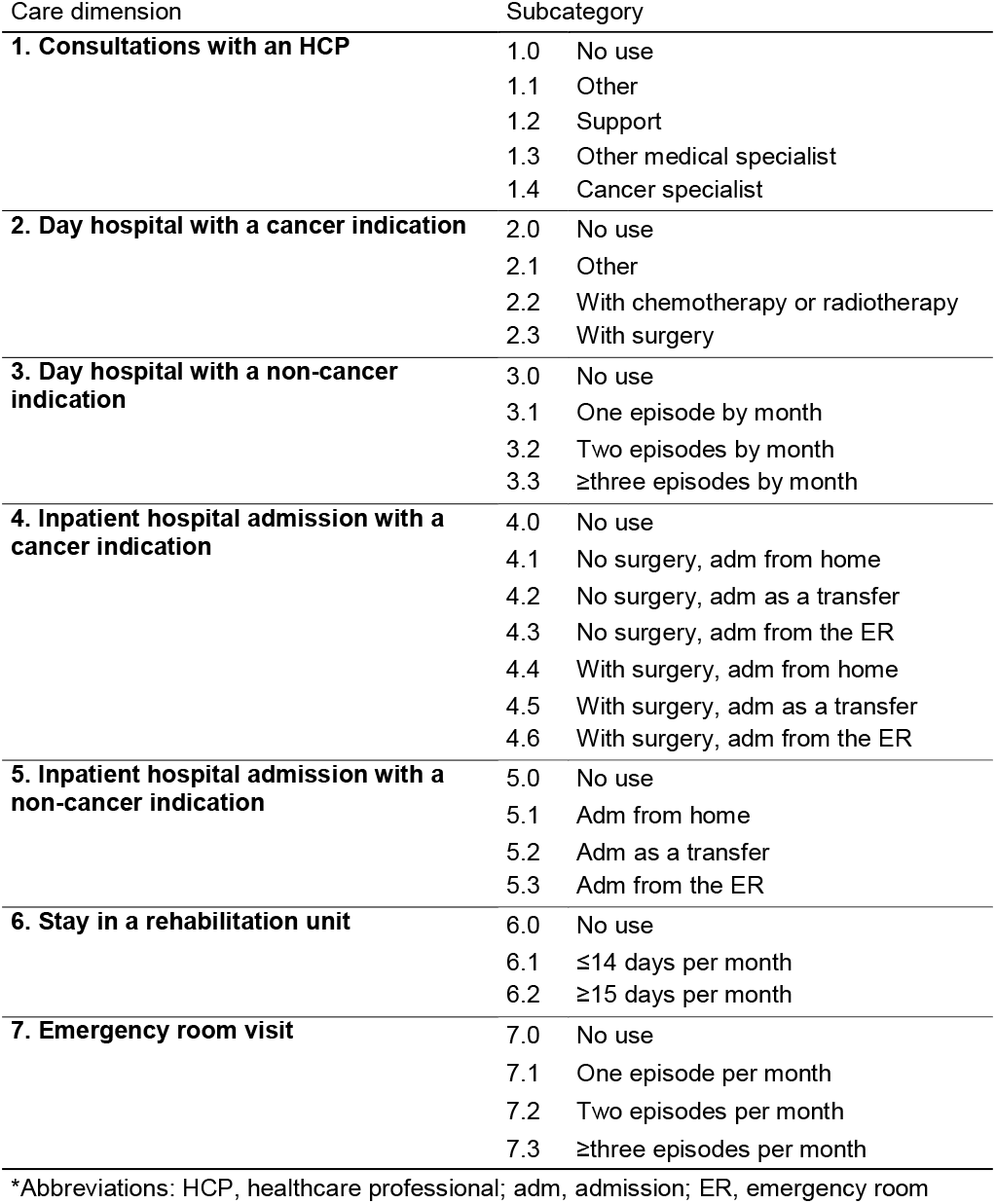
Hospital care dimensions.

#### Hospital care trajectory modelling

We retrospectively constructed individual “raw” hospital care trajectories from the chronological concatenation of all the care episodes. The individual trajectories were then modelled in a multichannel sequence analysis as a monthly aggregate of care consumption in each of the seven dimensions. For concomitant care episodes, only the highest-ranked subdimension was selected for each month and each dimension. Each patient’s hospital trajectories were then summarized into a 7 (dimensions) x 14 (months) matrix.

#### Clinical characteristics

The patients’ baseline clinical characteristics were extracted from the ELCAPA case report form. The variables included sociodemographic data, oncological parameters, a frailty screening tool (the G8 score (range: 0-17; impaired when ≤14/17)) and general and geriatric parameters (SM 3): Eastern Cooperative Oncology Group Performance Status (ECOG-PS, from 0 = normal to 4 = completely disabled), asthenia, mobility, autonomy, nutrition, cognition, mood, comorbidities, and polypharmacy. Lastly, the planned treatment was recorded, along with any scheduled surgical treatments, chemotherapy, and/or radiotherapy.

### Statistical analysis

A detailed description of the statistical analysis is given in SM 4. Briefly, clustering analysis relied on i) construction of a dissimilarity matrix representing the pairwise differences or ‘distances’ between patients using the optimal matching method, with an increased consideration given towards hospital admissions with surgery and cancer outpatient care and episode sequences rather than exact timing, to account for the clinical relevance of these aspects of care; ii) applying hierarchical clustering on this matrix to identify clusters of care trajectories using the Wards’s linkage criterion. Resulting clusters (the study’s primary endpoint) were then described and compared on patients’ geriatric and oncological characteristics, as well as on care consumption indicators.

To do so, we computed the total number of episodes per patient-month (ppm) of follow-up within each cluster, along with the numbers of planned and unplanned (based on the admission mode) episodes ppm. The hospital length of stay (LOS) overall and for planned and unplanned admissions was computed for each cluster by summing the number of days spent in hospital for inpatient and outpatient care (consultations and emergency room visits were therefore excluded) ppm of follow-up.

Multiple one-vs-rest multivariable logistic regression modeling was used to identify the best predictors of clusters belonging.

For all analyses, missing data were imputed using random forests trained on the observed values; the residual mean square error and the out-of-bag error were computed as a guide to imputation performance. Data were collected and preprocessed with Python 3.8 software and the AP-HP’s open-source libraries eds-scikit (18) and edsnlp (19). Data were imputed with the MissForest package (20), and care trajectories were modelled and clustered with the TraMineR (21) package in R 4.0 software (R Foundation, Vienna, Austria).

## Results

### Study population

As of March 31^st^, 2022, 6509 patients had been included in the ELCAPA study (SM 5). Of these, 4829 had been enrolled in an AP-HP center; 2903 of the latter had been enrolled between January 2012 and March 2019, and 2787 of the latter had a solid tumor.

During the database linkage process, we excluded 22 patients lacking an AP-HP identifier recorded in the ELCAPA database, 191 patients whose identifier was not found in the CDW, and 90 patients whose initials, date of birth or sex were not identical in the two databases. Next, we excluded 1168 correctly matched patients whose index date occurred before the CDW data completeness date, along with 608 patients with missing data or who had no care episodes recorded in the CDW during the study period. Hence, the study sample included 707 patients (median age: 82; range: 78-86), of whom 370 (52.3%) were male, 311 (45.2%) had a metastatic cancer, and 579 (86.3%) had an impaired G8 score. Relative to the nonincluded patients (n=2080, SM 6), the included patients had a significantly lower prevalence of metastatic cancer, a higher prevalence of curative treatment plans, and better cognitive status. Conversely, the included patients had a higher prevalence of polypharmacy.

A total of 10,998 care episodes [10,393 (94.5%) planned and 605 (5.5%) unplanned] were identified during the 14-month follow-up period; this corresponded to 4.7 days (95% confidence interval (CI) [4.65;4.76]) ppm spent in hospital overall, 3.82 [3.77;3.87] days ppm for planned episodes, and 0.89 [0.86;0.91] days ppm for unplanned episodes (Table 2).

**Table 2.**
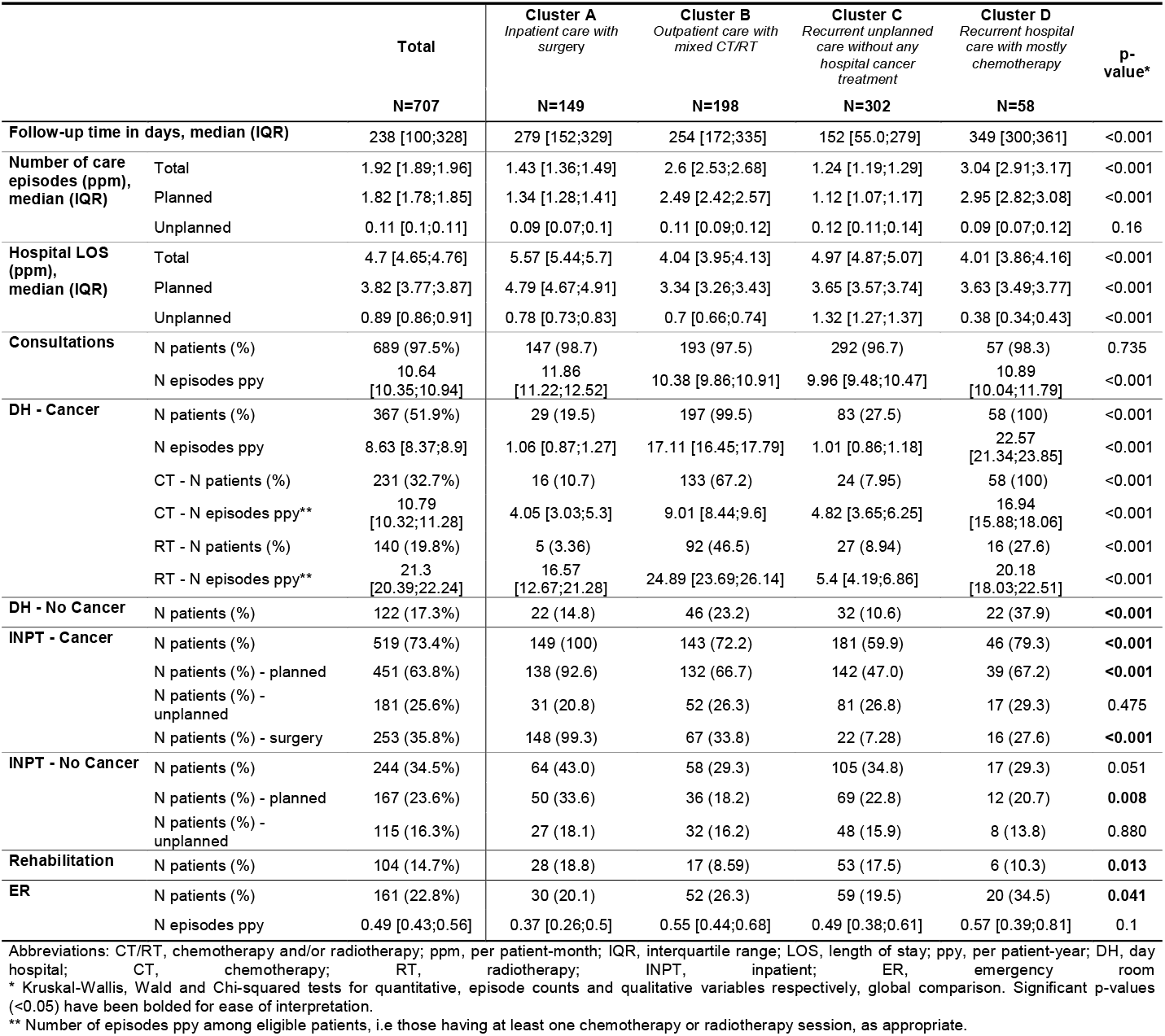
Care consumption indicators, overall and by cluster.

### Cluster analysis results and associated care consumption indicators

Four clusters of hospital care trajectories emerged from our analysis: in-hospital surgical trajectories (cluster A), mixed chemotherapy and/or radiotherapy (cluster B), absence of hospital cancer treatment (cluster C), and chemotherapy-dominant trajectories (cluster D). The clustering dendrogram and inertia curve are given in SM 7.

Cluster A (“*inpatient care with cancer surgery*”, n=149, 21.1%) comprised patients with a high level of consumption of inpatient care: 148 (99.3%) had undergone cancer surgery, 64 (43.0%) had been hospitalized for a non-cancer-related indication, and 28 (18.8%) had been admitted to a rehabilitation unit (Figure 1 and Table 2). Patients in this cluster had the longest LOS (5.57 [5.44;5.7] days ppm overall, with a median of 1.43 [1.36;1.49] care episodes ppm). The death rate in this cluster was 0.40 per patient-year (ppy); early death occurred for 14 patients (35.9% of the deaths in this cluster); 20 (51.3%) occurred in hospital, and 3 (15% of the latter) occurred after the provision of palliative care.

**Figure 1.**
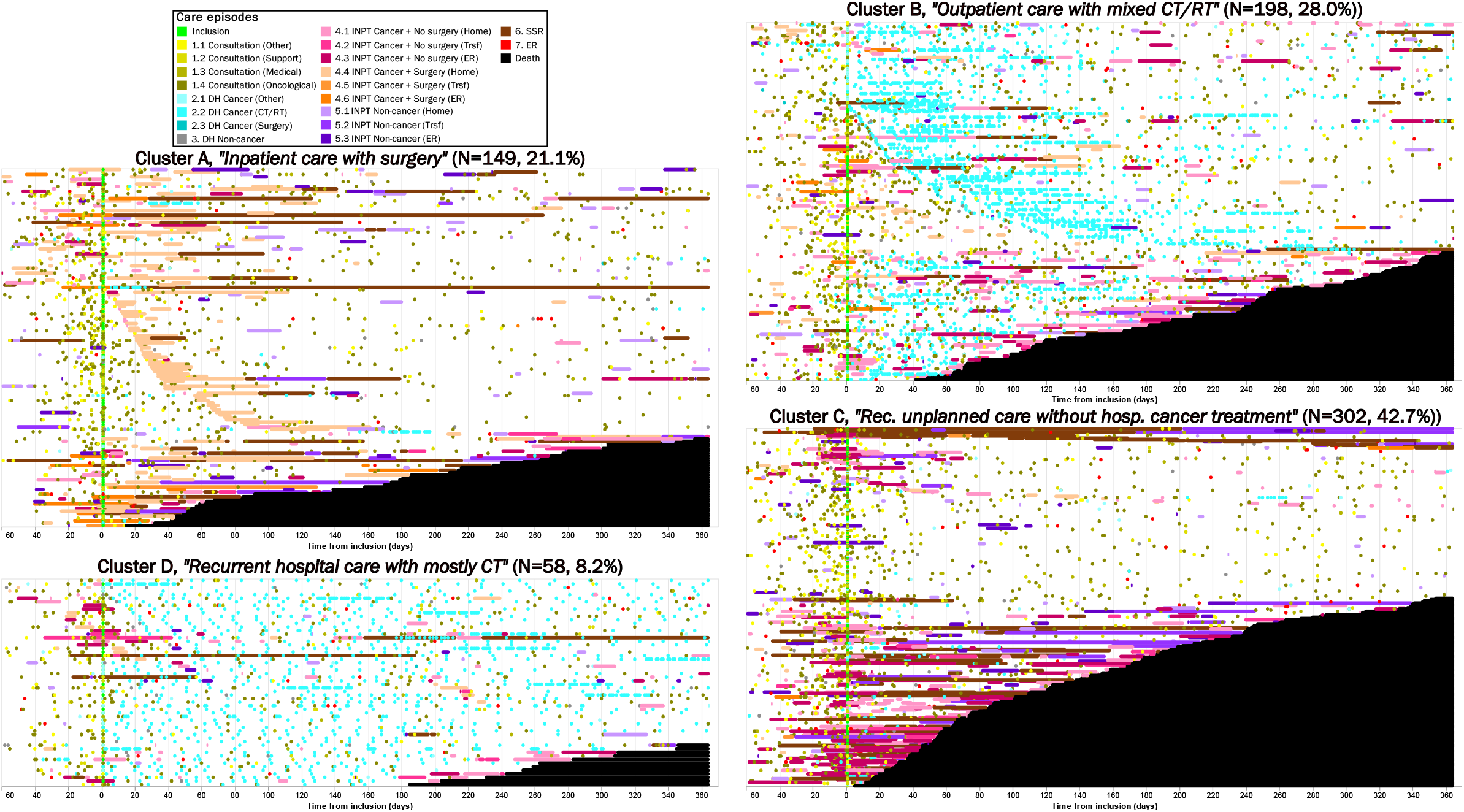
Chronograms of each cluster. Four clusters of hospital care trajectories are represented. Chronograms display the vertical concatenation of all the individual trajectories within each cluster, from 2 months before inclusion in ELCAPA (green dots) until 12 months after. Abbreviations: DH, day hospital; CT/RT, chemotherapy or radiotherapy; INPT, inpatient hospital admission; Trsf, transfer; ER, emergency room; Rehab, rehabilitation.

Conversely, most of the patients in cluster B (*“outpatient care with mixed chemotherapy and/or radiotherapy”*, n=198, 28.0%) received outpatient cancer care: the number of care episodes was 2.6 [2.53;2.68] ppm, and the LOS was short (4.04 [3.95;4.13] days ppm). 133 (67.2%) and 92 (46.5%) patients had at least one chemotherapy session or radiotherapy session, respectively. During follow-up, 0.59 deaths ppy occurred in this cluster, 15 (19.2%) were considered early; 41 (52.6%) deaths occurred in hospital, and 19 (46.3% of the latter) occurred after the provision of palliative care.

Cluster C was the largest cluster (*“recurrent unplanned care without any hospital cancer treatment”*, n=302, 42.7%) and comprised patients without hospital-specific cancer treatment procedures. The cluster’s members had the lowest number of care episodes ppm but the highest consumption of unplanned hospital care: they had 0.12 [0.11;0.14] unplanned episodes ppm and spent 1.32 [1.27;1.37] days ppm in hospital for unplanned episodes (4.97 [4.87;5.07] days ppm overall). This cluster had the highest 1-year death rate (1.22 deaths ppy), and the highest proportion of early deaths (80, 47.6% of the deaths in this cluster); 93 (55.4%) occurred in hospital, and 43 (46.2% of the latter) occurred after the provision of palliative care.

The 58 (8.2%) patients enrolled in cluster D (*“recurrent hospital care with mostly chemotherapy”*) had the highest number of care episodes ppm (mostly planned: 2.95 [2.82;3.08] ppm, and 3.04 [2.91;3.17] overall) and high levels of both inpatient and outpatient care consumption. All the patients received chemotherapy (16.94 [15.88;18.06] sessions ppy), and 46 (79.3%) patients had been hospitalized for cancer treatment or follow-up. They also had the highest proportion of non-cancer-related day-hospital visits, and the highest number of ER visits. This cluster had the lowest death rate during follow-up (0.23 deaths ppy), and no early death was recorded.

For illustrative purpose, we manually selected one patient from each cluster and described their individual hospital care trajectories in Figure 2. The sensitivity analysis of patients with newly diagnosed cancer gave clusters that were similar to those of the main analysis in terms of the patient distribution and phenotypes (results reported in SM 8).

**Figure 2.**
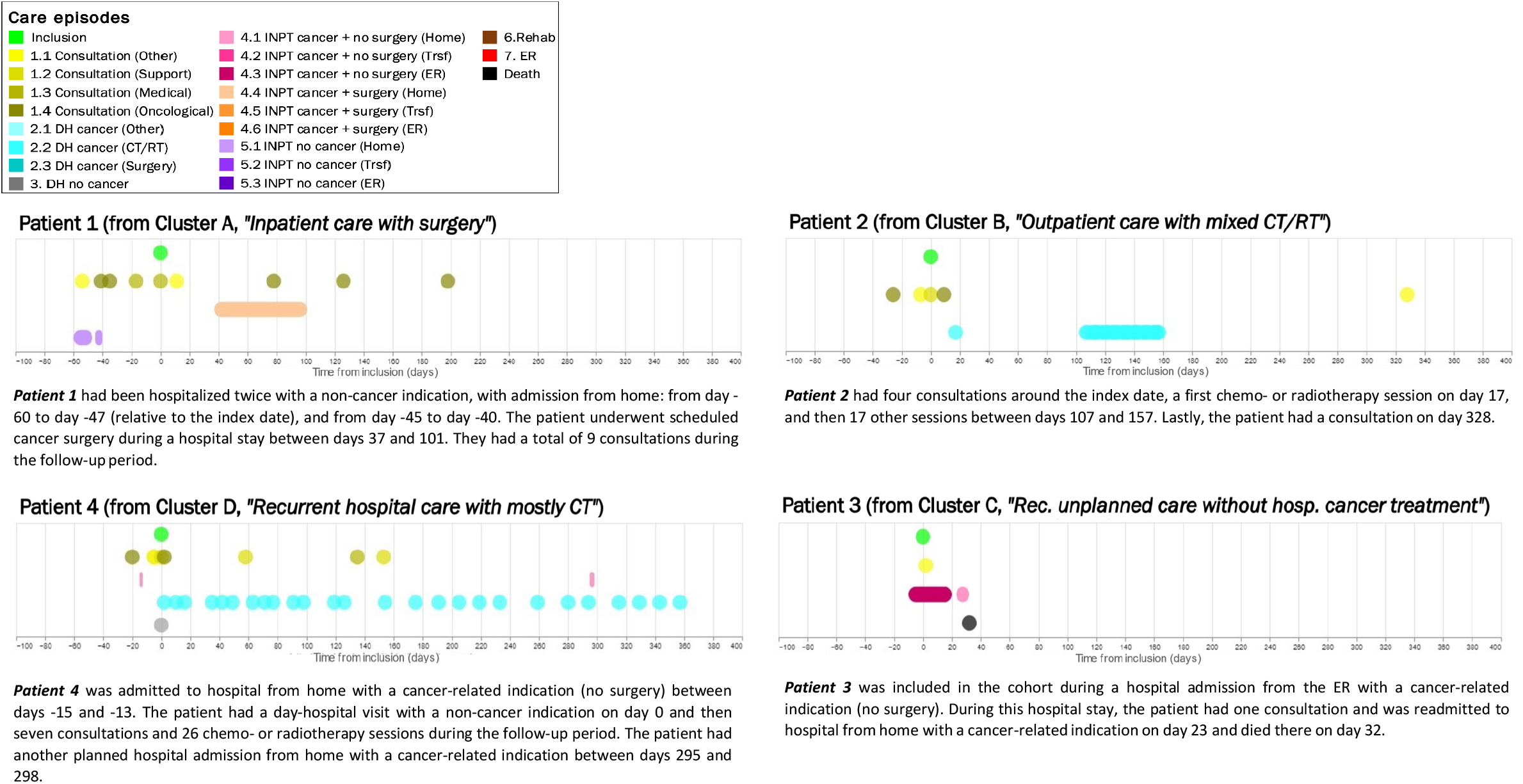
Selected individual patient trajectories from each cluster. Abbreviations: DH, day hospital; CT/RT, chemotherapy or radiotherapy; INPT, inpatient hospital admission; Trsf, transfer; ER, emergency room; Rehab, rehabilitation. Rec., recurrent; hosp., hospital.

### Geriatric and oncological parameters across clusters

Relative to other patients, those in cluster A (“*inpatient care with cancer surgery*”, median (range) age: 82 [78;86]; males: 57.0%) had a significantly lower proportion of metastatic cancer, a higher proportion of colorectal cancer and were in better general health (ECOG-PS score of 0 or 1) (Table 3).

**Table 3.**
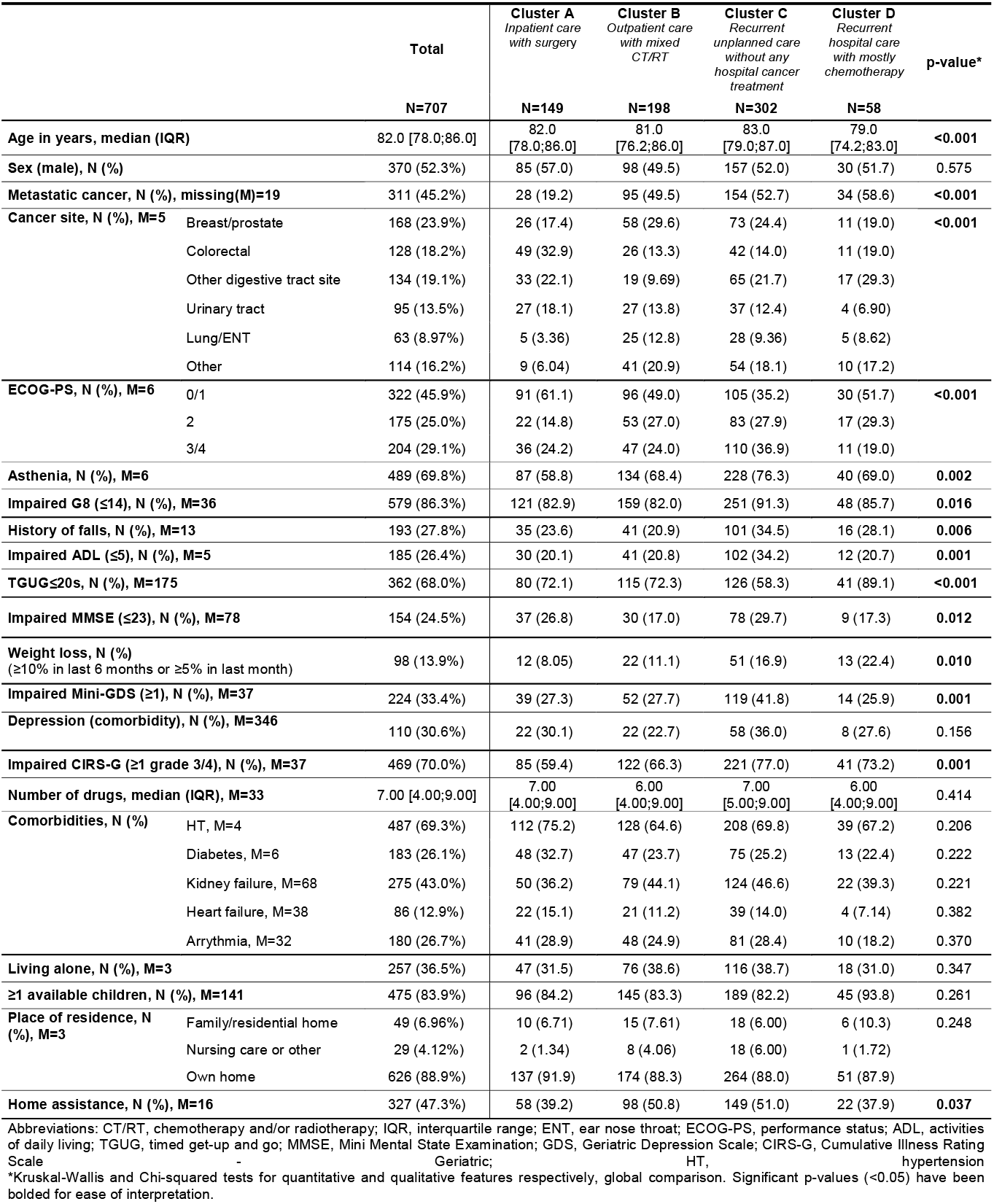
Clinical features, overall and by cluster.

Patients in cluster C (*“recurrent unplanned care without any hospital cancer treatment”*, median (range) age: 83 [79;87]; males: 52.0%) were most likely to have a poor ECOG-PS (score of 3 or 4) and impaired geriatric parameters (including impaired G8 score, ADL score, MMSE score, mini-GDS score, and CIRS-G score).

The patients in cluster B (“*outpatient care with mixed chemotherapy and/or radiotherapy”*) and D (*“recurrent hospital care with mostly chemotherapy*”) did not differ significantly with regard to clinical characteristics, except for age, TGUG status and cancer site: cluster D included the youngest patients, with the lowest proportion of impaired TGUG. Cluster B had a higher prevalence of breast/prostate, urinary tract and lung/ENT cancers, and cluster D had a higher prevalence of digestive cancers.

### Main factors predicting cluster membership

Significant interactions between cancer site and metastatic status as predictors of clusters B et C membership were found. An interaction variable was built accordingly and included in all regression models.

According to the one-vs-rest multivariable logistic regressions (Table 4), the factors associated independently with being in cluster A (“*inpatient care with cancer surgery*”) included having a localized digestive tract cancer (colorectal or other), a localized urinary tract cancer, an ECOG-PS score of 0 or 1, and hypertension. On the contrary, significant associations were found the absence of recent weight loss, the absence of kidney failure. Non-significant trends for the absence of asthenia and not living alone were noted.

**Table 4.** Cluster membership determinants.

|  |  | Cluster A |  |  | Cluster B |  |  | Cluster C |  |  | Cluster D |  |  |
| --- | --- | --- | --- | --- | --- | --- | --- | --- | --- | --- | --- | --- | --- |
|  |  | OR | 95% CI | p | OR | 95% CI | p | OR | 95% CI | p | OR | 95% CI | p |
| **Age** |  | - | - | - | - | - | - | 1.04 | [1.01-1.07] | **0.005** | 0.93 | [0.88-0.97] | **0.003** |
| **Cancer site** | **Metastatic cancer** |  |  |  |  |  |  |  |  |  |  |  |  |
| Breast/prostate | False | - | - | - | - | - | - | - | - | - | - | - | - |
|  | True | 0.56 | [0.21-1.37] | 0.218 | 0.79 | [0.40-1.53] | 0.479 | 1.87 | [0.97-3.63] | 0.061 | 0.74 | [0.18-2.61] | 0.647 |
| Colorectal | False | 5.94 | [2.91-12.5] | **<0.001** | 0.26 | [0.11-0.57] | **0.001** | 0.48 | [0.23-0.97] | **0.042** | 1.04 | [0.29-3.49] | 0.947 |
|  | True | 0.89 | [0.35-2.17] | 0.806 | 0.66 | [0.31-1.34] | 0.254 | 1.20 | [0.59-2.44] | 0.617 | 1.89 | [0.56-6.24] | 0.289 |
| Other digestive tract | False | 2.22 | [1.09-4.58] | **0.029** | 0.23 | [0.10-0.48] | **<0.001** | 1.41 | [0.75-2.65] | 0.284 | 1.18 | [0.39-3.58] | 0.769 |
|  | True | 0.42 | [0.11-1.24] | 0.145 | 0.32 | [0.12-0.73] | **0.010** | 2.56 | [1.23-5.43] | **0.013** | 2.40 | [0.81-7.39] | 0.116 |
| Lung/ENT | False | 0.58 | [0.13-1.98] | 0.425 | 1.03 | [0.39-2.64] | 0.951 | 1.97 | [0.78-5.04] | 0.153 | - | - | - |
|  | True | 0.24 | [0.04-0.93] | 0.071 | 1.28 | [0.57-2.84] | 0.541 | 1.07 | [0.46-2.45] | 0.866 | 1.46 | [0.39-5.10] | 0.559 |
| Urinary tract | False | 3.19 | [1.46-7.08] | **0.004** | 0.50 | [0.22-1.05] | 0.073 | 1.01 | [0.49-2.09] | 0.969 | 0.00 | [0.00-0.00] | 0.985 |
|  | True | 0.58 | [0.15-1.77] | 0.372 | 0.95 | [0.42-2.10] | 0.901 | 1.56 | [0.70-3.43] | 0.272 | 1.27 | [0.31-4.61] | 0.723 |
| Other | False | 0.57 | [0.19-1.51] | 0.280 | 1.78 | [0.87-3.65] | 0.117 | 0.75 | [0.36-1.57] | 0.455 | 1.32 | [0.33-4.76] | 0.674 |
|  | True | 0.26 | [0.06-0.84] | **0.041** | 0.69 | [0.33-1.40] | 0.304 | 2.03 | [1.02-4.10] | **0.046** | 1.44 | [0.43-4.68] | 0.543 |
| **ECOG-PS** | 0/1 | - | - | - | - | - | - | - | - | - | - | - | - |
|  | 2 | 0.56 | [0.31;0.98] | **0.048** | - | - | - | - | - | - | - | - | - |
|  | 3/4 | 0.89 | [0.53;1.49] | 0.668 | - | - | - | - | - | - | - | - | - |
| **Asthenia** |  | 0.64 | [0.41-1.00] | 0.051 | - | - | - | - | - | - | - | - | - |
| **History of falls** |  | - | - | - | 0.62 | [0.40-0.94] | **0.025** | 1.42 | [0.99-2.03] | 0.059 | - | - | - |
| **TGUG≤20s** |  | - | - | - | - | - | - | 0.63 | [0.45-0.89] | **0.009** | 2.62 | [1.36-5.48] | **0.006** |
| **MMSE score** |  | - | - | - | 1.10 | [1.05-1.16] | **<0.001** | 0.94 | [0.91-0.98] | **0.005** | - | - | - |
| **Weight loss** |  | 0.43 | [0.20-0.86] | **0.021** | - | - | - | 1.59 | [1.00-2.52] | 0.050 | 1.62 | [0.78-3.18] | 0.178 |
| **Impaired Mini-GDS** |  | - | - | - | - | - | - | 1.59 | [1.12-2.24] | **0.009** | - | - | - |
| **Depression (comorbidity)** |  | - | - | - | 0.69 | [0.45-1.03] | 0.073 | - | - | - | - | - | - |
| **Comorbidities** | HT | 1.64 | [1.04-2.63] | **0.038** | 0.75 | [0.52-1.09] | 0.131 | - | - | - | - | - | - |
|  | Kidney failure | 0.63 | [0.40-0.97] | 0.038 | - | - | - | - | - | - | - | - | - |
| **Living alone** |  | 0.64 | [0.41-1.00] | 0.050 | - | - | - | - | - | - | - | - | - |
| **Place of residence** | Own home | - | - | - | - | - | - | - | - | - | - | - | - |
|  | Family/residential home | 1.51 | [0.65-3.26] | 0.312 | - | - | - | - | - | - | - | - | - |
|  | Nursing care or other | 0.21 | [0.03-0.85] | 0.056 | - | - | - | - | - | - | - | - | - |
| **Home assistance** |  | - | - | - | 1.48 | [1.04-2.12] | **0.032** | - | - | - | - | - | - |
| **≥1 Available children** |  | - | - | - | - | - | - | - | - | - | 2.82 | [0.98-11.9] | **0.09** |

Abbreviations: OR, odds ratio; CI, confidence interval; p, p-value; ENT, ear nose throat; ECOG-PS, performance status; TGUG, timed get-up-and-go; MMSE, Mini Mental State Examination; GDS, Geriatric Depression Scale; HT, hypertension

The factors associated independently with membership of clusters B (“*outpatient care with mixed chemotherapy and/or radiotherapy”*) or C (*“recurrent unplanned care without any hospital cancer treatment”*) were related to geriatric parameters. Patients with a higher MMSE score, no history of falls, and requiring home assistance were more likely to be in cluster B. Significant associations with the absence of a localized colorectal cancer and other digestive tract cancer were also found in cluster B. On the contrary, patients with impaired mobility (TGUG>20s), a lower MMSE score, an impaired mini-GDS score, older age and a digestive metastatic cancer were more likely to be included in cluster C. In this cluster, non-significant trends were observed for metastatic breast or prostate cancer, history of falls and recent weight loss.

Lastly, the factors associated independently with cluster D membership (*“recurrent hospital care with mostly chemotherapy”*) included younger age and having a TGUG ≤ 20s. Nonsignificant trend for having at least one child able to provide support was also noted.

Cluster C’s chronogram could also be split visually into two subgroups by death status, each having specific hospital care consumption patterns and associated clinical profiles (SM 9).

## Discussion

We identified four hospital care trajectory clusters among a population of 70 years or older patients with solid cancer: in-hospital surgical trajectories (cluster A), outpatient care trajectories with mixed chemotherapy and/or radiotherapy (cluster B), absence of hospital cancer treatment (cluster C) and chemotherapy-dominant with high hospital care consumption trajectories (cluster D). We did not find any associations between surgical trajectories on one hand and geriatric parameters or age on the other. However, we found that geriatric parameters were independently associated with cluster B when unimpaired and with cluster C when impaired. Younger age was associated with chemotherapy-dominant trajectories. Lastly, by further exploring cluster C, we identified two subgroups of trajectories: (i) frequent, prolonged, and unplanned care consumption until early death, and (ii) steady consultations during the follow-up period. Both of the latter were associated with specific clinical profiles.

To our knowledge, only one study by Depoorter et al.(22) has reported on the hospital care consumption of older adults with cancer. They found smaller care consumption indices than in our study (0.43 vs 0.49 ER visits ppy, 11.86 vs 56.4 days ppy spent in hospital). This difference may be attributed to the advanced frailty status and cancer severity in our cohort (64.3% of the patients included in their cohort had an impaired G8 score, compared to 86.3% in this study; and 16.8% vs 45.2% had metastatic cancer), and to their decision to exclude end-of-life care (which is included in this study), known to be resource-intensive. Notably, they did not consider individual trajectories (except for living situations), nor did they further characterize the hospital care consumption with regards to the associated patients’ profiles. Other studies applied automatic clustering to claims data from all-age patients with specific tumor sites and treatment settings, and found similar patterns of health care utilization (7, 23, 24). The absence of associations between age and geriatric parameters and cluster A (“*inpatient care with cancer surgery*”) in the present study suggests that patients, despite having older age and/or geriatric impairments, are still offered surgical opportunities to treat their cancer. The long hospital LOS and the high proportion of patients receiving post-surgery rehabilitation care suggests that the care trajectories were well planned. Conversely, impaired mobility, cognition, and mood status were independently associated with low-intensity hospital cancer treatment. This finding is in line with previous research. For example, Caillet et al. found that functional impairment and malnutrition were associated with lower-intensity cancer treatment(13), and Sourdet et al. reported similar associations with cognitive, nutritional and mobility parameters (25). These geriatric impairments are also known to be associated with unplanned hospital admissions (26-28), which is related to the care consumption observed for patients in cluster C (“*recurrent unplanned care without any hospital cancer treatment”*). Interestingly, our results highlighted a significant association between younger age and cluster D membership (*“recurrent hospital care with mostly chemotherapy”*). This suggests that compliance with the current guidelines promoting decisions based on functional age (defined as a combination of physiological, psychological and social age, in opposition to chronological, *i*.*e*. calendar age) may be difficult in a real-world setting for chemotherapy (29-32).

Our results suggest two implications for decision-making. Firstly, our analysis provided useful insights on the real-world experiences of older patients with cancer to help defining evidence-based clinical pathways adapted to this specific population. Secondly, our analysis highlighted the requirement for dedicated in-hospital facilities for high-need patients who have the worst prognosis (cluster C): these patients may not be the first beneficiaries of both rehabilitation care (because no treatments are scheduled) and end-of-lifecare (because they are not at risk of imminent death), and they do not seem to fit into any of today’s care pathways.

Our study had a number of limitations. We retrieved data from the EHRs of 39 public-sector hospitals, which together account for only one third of the hospital beds in acute wards available in the Greater Paris Area. This limited the exhaustiveness of our healthcare trajectory reconstruction, and might have underestimated the hospital care consumption of patients. Furthermore, not all types of cancer treatment were documented, especially treatment with oral chemotherapy drugs and radiotherapy sessions in private-sector establishments. Finally, our study setting involved a geriatric assessment (GA) for all patients prior to a decision on cancer treatment. Hopefully, subsequent hospital care trajectories should be impacted by the GA results (although this might require formal proof), and performing the same clustering analysis in a clinical setting without systematic GA might lead to different results and interpretations.

Our study had a number of strengths. The linkage of clinical and medical administrative databases provided insights into care consumption and thorough clinical phenotypes, which have often been missing in previous studies of care trajectories (10-12, 33). Furthermore, our custom-based dissimilarity metric highlighting the main hospital cancer treatment procedures enabled us to build clinically meaningful clusters of trajectories. Lastly, we were able to identify high-need (and probably high-cost) patients at risk of an adverse trajectory and who probably require better healthcare planning. Future work should focus on exploring the hospital care trajectories of patients at a disease-specific level, to identify health and fitness thresholds that have a direct clinical application.

## Conclusion

By linking a cohort study database and a CDW, we identified four clusters of care trajectories among older patients with solid cancer. The clusters differed primarily with regard to their main cancer treatment procedures. We did not find any significant associations between geriatric parameters and surgical trajectories. Conversely, patients with impairments in geriatric parameters were associated with trajectories that lacked hospital cancer treatment; this subgroup of high-need patients requires better health care planning towards the end of their life. Lastly, we found that younger age was independently associated with chemotherapy-dominant trajectories.

## Additional Information

## Acknowledgments

ELCAPA Study Group comprises geriatricians (Amelie Aregui, Geoffroy Beraud Chaulet, Mickaël Bringuier, Philippe Caillet, Pascale Codis, Lola Corsin, Tristan Cudennec, Anne Chahwakilian, Amina Djender, Virginie Fossey-Diaz, Mathilde Gisselbrecht, Betarice Gonzalez,Aissata Konate, Marie Laurent, Madeleine Lefevre, Emmanuelle Lorisson,Julien Le Guen Josephine Massias, Soraya Merbaki,Galdric Orvoen, Elena Paillaud, Johanne Poisson, Frédéric Pamoukdjian, Anne-Laure Scain, Godelieve Rochette de Lempdes, Caryn Recto,Florence Rollot-Trad, Kevin Rougette, Umay Saritayli, Zoe Ap Thomas,Selim Turk, Hélène Vincent, Johanna Vouriot), oncologists (Pascaline Boudou-Rouquette,Marc-Antoine Benderra, Stéphane Culine, Etienne Brain, Maxime Frelaut, Alexandre de Moura, Aurelien Noret,Romain Geiss, and Christophe Tournigand), a digestive oncologist (Thomas Aparicio), a gynecological oncologist (Cyril Touboul), a radiation oncologist (Jean-Léon Lagrange), epidemiologists (Etienne Audureau, Sylvie Bastuji-Garin and Florence Canouï-Poitrine), a medical biologist (Marie-Anne Loriot), a pharmacist (Pierre-André Natella), a biostatistician (Claudia Martinez-Tapia), a clinical research physician (Nicoleta Reinald), a clinical research nurse (Florence Devillard), a datamanager (Clélia Chambraud), and clinical research assistants (Aurélie Baudin, Sabrina Chaoui, Johanna Canovas, Axelle Histe, Sandrine Lacour,Laure Morisset, Besma Saasaoui, Debhbia Yachir,).

Data used in preparation of this article were obtained from the AP-HP Clinical Data Warehouse (CDW). As such, members of the AP-HP CDW contributed to the design and implementation of the database, and participated in the databases linkage process. A complete list of the AP-HP CDW members can be found at https://eds.aphp.fr/.

The authors wish to thank David Fraser, PhD, for proofreading the manuscript.

## Data Availability

Access to the clinical data warehouse’s de-identified raw data can be granted following the process described on its website: www.eds.aphp.fr. A prior validation of the access by the local institutional review board is required. In the case of non-AP-HP researchers, the signature of a collaboration contract is moreover mandatory.

## References

1. Cancer today [Available from: https://gco.iarc.fr/today/online-analysis-table?v=2020&mode=population&mode_population=who&population=900&populations=900&key=asr&sex=0&cancer=39&type=0&statistic=5&prevalence=0&population_group=0&ages_group%5B%5D=0&ages_group%5B%5D=17&group_cancer=1&include_nmsc=0&include_nmsc_other=1.

2. Administration USDoHaHSFaD. Inclusion of Older Adults in Cancer Clinical Trials. 2022.

3. Javier-DesLoges J, Nelson TJ, Murphy JD, McKay RR, Pan E, Parsons JK, et al. Disparities and trends in the participation of minorities, women, and the elderly in breast, colorectal, lung, and prostate cancer clinical trials. Cancer. 2022;128(4):770–7.

4. Pitkala KH, Strandberg TE. Clinical trials in older people. Age Ageing. 2022;51(5).

5. Todd OM, Burton JK, Dodds RM, Hollinghurst J, Lyons RA, Quinn TJ, et al. New Horizons in the use of routine data for ageing research. Age Ageing. 2020;49(5):716–22.

6. Hamaker ME, Te Molder M, Thielen N, van Munster BC, Schiphorst AH, van Huis LH. The effect of a geriatric evaluation on treatment decisions and outcome for older cancer patients - A systematic review. J Geriatr Oncol. 2018;9(5):430–40.

7. Zhuang Q, Chong PH, Ong WS, Yeo ZZ, Foo CQZ, Yap SY, et al. Longitudinal patterns and predictors of healthcare utilization among cancer patients on home-based palliative care in Singapore: a group-based multi-trajectory analysis. BMC Med. 2022;20(1):313.

8. Langton JM, Reeve R, Srasuebkul P, Haas M, Viney R, Currow D, et al. Health service use and costs in the last 6 months of life in elderly decedents with a history of cancer: a comprehensive analysis from a health payer perspective. Br J Cancer. 2016;114(11):1293–302.

9. Luta X, Diernberger K, Bowden J, Droney J, Hall P, Marti J. Intensity of care in cancer patients in the last year of life: a retrospective data linkage study. Br J Cancer. 2022;127(4):712–9.

10. Elyn A, Gardette V, Renoux A, Sourdet S, Nourhashemi F, Sanou B, et al. Potential determinants of unfavourable healthcare utilisation trajectories during the last year of life of people with incident Alzheimer Disease or Related Syndromes: a nationwide cohort study using administrative data. Age Ageing. 2022;51(3).

11. Vanasse A, Courteau J, Courteau M, Benigeri M, Chiu YM, Dufour I, et al. Healthcare utilization after a first hospitalization for COPD: a new approach of State Sequence Analysis based on the ‘6W’ multidimensional model of care trajectories. BMC Health Serv Res. 2020;20(1):177.

12. Tsai TH, Huang N, Lin IF, Chou YJ. Variation in the 11-year trajectories of medical care seeking behaviors in diabetes patients under a single payer system: persisting gaps to be filled. BMC Health Serv Res. 2019;19(1):580.

13. Caillet P, Canoui-Poitrine F, Vouriot J, Berle M, Reinald N, Krypciak S, et al. Comprehensive geriatric assessment in the decision-making process in elderly patients with cancer: ELCAPA study. J Clin Oncol. 2011;29(27):3636–42.

14. Torres MJ, Coste J, Canoui-Poitrine F, Pouchot J, Rachas A, Carcaillon-Bentata L. Impact of the first COVID-19 pandemic wave on hospitalizations and deaths caused by geriatric syndromes in France: a nationwide study. J Gerontol A Biol Sci Med Sci. 2023.

15. Boulahssass R, Gonfrier S, Ferrero JM, Sanchez M, Mari V, Moranne O, et al. Predicting early death in older adults with cancer. Eur J Cancer. 2018;100:65–74.

16. Rosen AK, Mayer-Oakes A. Episodes of care: theoretical frameworks versus current operational realities. Jt Comm J Qual Improv. 1999;25(3):111–28.

17. France SP. Algorithme de sélection des hospitalisations liées au cancer en MCO /étude de validation. In: INCa, editor. 2018.

18. Petit-Jean TR A.; Maladière, V.; Varoquaux, G.; Bey, R. eds-scikit: data analysis on OMOP databases.

19. Dura BW, P.; Petit-Jean, T.; Cohen, A.; Jean, C.; Bey, R. EDS-NLP: efficient information extraction from French clinical notes.

20. Stekhoven DJ. missForest: Nonparametric Missing Value Imputation using Random Forest. 2022.

21. Gabadinho AS, M.; Müller, N.; Bürgin, R.; Fonta, P.A.; Ritschard, G. Trajectory Miner: a Toolbox for Exploring and Rendering Sequences. 2023.

22. Depoorter V, Vanschoenbeek K, Decoster L, Silversmit G, Debruyne PR, De Groof I, et al. Long-term health-care utilisation in older patients with cancer and the association with the Geriatric 8 screening tool: a retrospective analysis using linked clinical and population-based data in Belgium. Lancet Healthy Longev. 2023;4(7):e326–e36.

23. Baudrier C, Tran Y, Delanoy N, Katsahian S, Sabatier B, Perrin G. Identifying homogeneous healthcare use profiles and treatment sequences by combining sequence pattern mining with care trajectory clustering in kidney cancer patients on oral anticancer drugs: A case study. Health Informatics J. 2022;28(2):14604582221101526.

24. Schuler MS, Joyce NR, Huskamp HA, Lamont EB, Hatfield LA. Medicare Beneficiaries With Advanced Lung Cancer Experience Diverse Patterns Of Care From Diagnosis To Death. Health Aff (Millwood). 2017;36(7):1193–200.

25. Sourdet S, Brechemier D, Steinmeyer Z, Gerard S, Balardy L. Impact of the comprehensive geriatric assessment on treatment decision in geriatric oncology. BMC Cancer. 2020;20(1):384.

26. Feliu J, Espinosa E, Basterretxea L, Paredero I, Llabres E, Jimenez-Munarriz B, et al. Prediction of Unplanned Hospitalizations in Older Patients Treated with Chemotherapy. Cancers (Basel). 2021;13(6).

27. Lodewijckx E, Kenis C, Flamaing J, Debruyne P, De Groof I, Focan C, et al. Unplanned hospitalizations in older patients with cancer: Occurrence and predictive factors. J Geriatr Oncol. 2021;12(3):368–74.

28. Chiang LY, Liu J, Flood KL, Carroll MB, Piccirillo JF, Stark S, et al. Geriatric assessment as predictors of hospital readmission in older adults with cancer. J Geriatr Oncol. 2015;6(4):254–61.

29. Zon RT, Frame JN, Neuss MN, Page RD, Wollins DS, Stranne S, et al. American Society of Clinical Oncology Policy Statement on Clinical Pathways in Oncology. J Oncol Pract. 2016;12(3):261–6.

30. Meresse M, Bouhnik AD, Bendiane MK, Retornaz F, Rousseau F, Rey D, et al. Chemotherapy in Old Women with Breast Cancer: Is Age Still a Predictor for Under Treatment? Breast J. 2017;23(3):256–66.

31. Neal D, Morgan JL, Kenny R, Ormerod T, Reed MW. Is there evidence of age bias in breast cancer health care professionals’ treatment of older patients? Eur J Surg Oncol. 2022;48(12):2401–7.

32. Wildiers H, Heeren P, Puts M, Topinkova E, Janssen-Heijnen ML, Extermann M, et al. International Society of Geriatric Oncology consensus on geriatric assessment in older patients with cancer. J Clin Oncol. 2014;32(24):2595–603.

33. Chouaid C, Grumberg V, Batisse A, Corre R, Giaj Levra M, Gaudin AF, et al. Machine Learning-Based Analysis of Treatment Sequences Typology in Advanced Non-Small-Cell Lung Cancer Long-Term Survivors Treated With Nivolumab. JCO Clin Cancer Inform. 2022;6:e2100108.

