## Supplementary Materials for "Hospital Care Trajectories of Older Adults with Cancer and the Associated Clinical Profiles: linking a French Prospective Cohort Study and a Clinical Data Warehouse"

### Supplementary material

Table of content

### SM 1. Chronological analysis of data completeness in the AP-HP’s CDW

The data in the AP-HP’s CDW goes back to 2012, and the volume has increased with the progressive implementation of shared EHR software across the group’s hospitals. To ensure data completeness in our analysis, we visualized the change over time in the volume of available CDW data for each of the AP-HP’s hospitals. We next fitted step functions to these time series, in order to define the data completeness dates by hospital. The dates were then aggregated by hospital group to give our study’s start date.

Results - The chronological analysis of data completeness across the AP-HP’s six hospital groups yielded dates ranging from October 2015 (for the *Paris-Saclay* group) to October 2017 (for the *Centre Paris Cité* group).

### SM 2. RECORD reporting guidelines

**The RECORD statement – checklist of items, extended from the STROBE statement, that should be reported in observational studies using routinely collected health data.**

|  | **Item No.** | **STROBE items** | **Location in manuscript where items are reported** | **RECORD items** | **Location in manuscript where items are reported** |
| --- | --- | --- | --- | --- | --- |
| **Title and abstract** | | | | | |
|  | 1 | (a) Indicate the study’s design with a commonly used term in the title or the abstract (b) Provide in the abstract an informative and balanced summary of what was done and what was found | (a) Abstract – *Patients and Methods*  (b) Abstract – *Patients and Methods* & *Results* | RECORD 1.1: The type of data used should be specified in the title or abstract. When possible, the name of the databases used should be included.  RECORD 1.2: If applicable, the geographic region and timeframe within which the study took place should be reported in the title or abstract.  RECORD 1.3: If linkage between databases was conducted for the study, this should be clearly stated in the title or abstract. | 1.1 Title and Abstract – *Patients and Methods* 1.2 Abstract – *Patients and Methods* 1.3 Title and Abstract – *Patients and Methods* |
| **Introduction** | | | | | |
| Background rationale | 2 | Explain the scientific background and rationale for the investigation being reported | Introduction (1^st^ paragraph) |  |  |
| Objectives | 3 | State specific objectives, including any prespecified hypotheses | Introduction (2^nd^ paragraph) |  |  |
| **Methods** | | | | | |
| Study Design | 4 | Present key elements of study design early in the paper | Methods – *Data sources, study design and setting* |  |  |
| Setting | 5 | Describe the setting, locations, and relevant dates, including periods of recruitment, exposure, follow-up, and data collection | Methods – *Data sources, study design and setting* |  |  |
| Participants | 6 | *(a) Cohort study* - Give the eligibility criteria, and the sources and methods of selection of participants. Describe methods of follow-up  *Case-control study* - Give the eligibility criteria, and the sources and methods of case ascertainment and control selection. Give the rationale for the choice of cases and controls  *Cross-sectional study* - Give the eligibility criteria, and the sources and methods of selection of participants  *(b) Cohort study* - For matched studies, give matching criteria and number of exposed and unexposed  *Case-control study* - For matched studies, give matching criteria and the number of controls per case | (a) Methods – *Study Population* | RECORD 6.1: The methods of study population selection (such as codes or algorithms used to identify subjects) should be listed in detail. If this is not possible, an explanation should be provided.  RECORD 6.2: Any validation studies of the codes or algorithms used to select the population should be referenced. If validation was conducted for this study and not published elsewhere, detailed methods and results should be provided.  RECORD 6.3: If the study involved linkage of databases, consider use of a flow diagram or other graphical display to demonstrate the data linkage process, including the number of individuals with linked data at each stage. | 6.1 Methods – *Study Population*  6.2 Methods – *Study Population*  6.3 Figure 1 |
| Variables | 7 | Clearly define all outcomes, exposures, predictors, potential confounders, and effect modifiers. Give diagnostic criteria, if applicable. | Methods – *Data collection*  Supplementary Materials 3 | RECORD 7.1: A complete list of codes and algorithms used to classify exposures, outcomes, confounders, and effect modifiers should be provided. If these cannot be reported, an explanation should be provided. | Methods – *Data collection*  Supplementary Materials 3 |
| Data sources/ measurement | 8 | For each variable of interest, give sources of data and details of methods of assessment (measurement).  Describe comparability of assessment methods if there is more than one group | Methods – *Data collection*  Supplementary Materials 1, 3 and 4 |  |  |
| Bias | 9 | Describe any efforts to address potential sources of bias | Methods – *Study Population* |  |  |
| Study size | 10 | Explain how the study size was arrived at | Methods – *Study Population and Data collection* |  |  |
| Quantitative variables | 11 | Explain how quantitative variables were handled in the analyses. If applicable, describe which groupings were chosen, and why | Methods – *Statistical analysis* |  |  |
| Statistical methods | 12 | (a) Describe all statistical methods, including those used to control for confounding  (b) Describe any methods used to examine subgroups and interactions  (c) Explain how missing data were addressed  (d) *Cohort study* - If applicable, explain how loss to follow-up was addressed  *Case-control study* - If applicable, explain how matching of cases and controls was addressed  *Cross-sectional study* - If applicable, describe analytical methods taking account of sampling strategy  (e) Describe any sensitivity analyses | (a), (b) and (c) Methods – *Statistical analysis*  (d) NA  (e) Methods – *Study Population* |  |  |
| Data access and cleaning methods |  | .. |  | RECORD 12.1: Authors should describe the extent to which the investigators had access to the database population used to create the study population.  RECORD 12.2: Authors should provide information on the data cleaning methods used in the study. | 12.1 Methods – *Data sources, study design and setting*  12.2 Methods – *Data sources, study design and setting* & *Data collection* & *Statistical analysis* |
| Linkage |  | .. |  | RECORD 12.3: State whether the study included person-level, institutional-level, or other data linkage across two or more databases. The methods of linkage and methods of linkage quality evaluation should be provided. | Methods - *Data sources, study design and setting* & *Study Population* |
| **Results** | | | | | |
| Participants | 13 | (a) Report the numbers of individuals at each stage of the study (*e.g.*, numbers potentially eligible, examined for eligibility, confirmed eligible, included in the study, completing follow-up, and analysed)  (b) Give reasons for non-participation at each stage.  (c) Consider use of a flow diagram | (a), (b) and (c) Results (1^st^ and 2^nd^ paragraphs), Figure 1 and SM 1 | RECORD 13.1: Describe in detail the selection of the persons included in the study (*i.e.,* study population selection) including filtering based on data quality, data availability and linkage. The selection of included persons can be described in the text and/or by means of the study flow diagram. | Results (1^st^ and 2^nd^ paragraphs), Figure 1 and SM 1 |
| Descriptive data | 14 | (a) Give characteristics of study participants (*e.g.*, demographic, clinical, social) and information on exposures and potential confounders  (b) Indicate the number of participants with missing data for each variable of interest  (c) *Cohort study* - summarise follow-up time (*e.g.*, average and total amount) | (a) Results (1^st^ paragraph), Table 2  (b) and (c) Table 2 |  |  |
| Outcome data | 15 | *Cohort study* - Report numbers of outcome events or summary measures over time  *Case-control study* - Report numbers in each exposure category, or summary measures of exposure  *Cross-sectional study* - Report numbers of outcome events or summary measures | Results (3^rd^ and 4^th^ paragraphs) and Table 2 |  |  |
| Main results | 16 | (a) Give unadjusted estimates and, if applicable, confounder-adjusted estimates and their precision (e.g., 95% confidence interval). Make clear which confounders were adjusted for and why they were included  (b) Report category boundaries when continuous variables were categorized  (c) If relevant, consider translating estimates of relative risk into absolute risk for a meaningful time period | (a) Results (13^th^, 14^th^ and 15^th^ paragraphs) and Table 2  (b) SM 3  (c) NA |  |  |
| Other analyses | 17 | Report other analyses done—e.g., analyses of subgroups and interactions, and sensitivity analyses | Results (9^th^ paragraph) |  |  |
| **Discussion** | | | | | |
| Key results | 18 | Summarise key results with reference to study objectives | Discussion (1^st^ paragraph) |  |  |
| Limitations | 19 | Discuss limitations of the study, taking into account sources of potential bias or imprecision. Discuss both direction and magnitude of any potential bias | Discussion (4^th^ paragraph) | RECORD 19.1: Discuss the implications of using data that were not created or collected to answer the specific research question(s). Include discussion of misclassification bias, unmeasured confounding, missing data, and changing eligibility over time, as they pertain to the study being reported. | Discussion (4^th^ paragraph) |
| Interpretation | 20 | Give a cautious overall interpretation of results considering objectives, limitations, multiplicity of analyses, results from similar studies, and other relevant evidence | Discussion (2^nd^ paragraph) |  |  |
| Generalisability | 21 | Discuss the generalisability (external validity) of the study results | Discussion (3^rd^ paragraph) |  |  |
| **Other Information** | | | | | |
| Funding | 22 | Give the source of funding and the role of the funders for the present study and, if applicable, for the original study on which the present article is based |  |  |  |
| Accessibility of protocol, raw data, and programming code |  | .. |  | RECORD 22.1: Authors should provide information on how to access any supplemental information such as the study protocol, raw data, or programming code. | Methods - *Data sources, study design and setting* & *Statistical analysis* |

*Reference: Benchimol EI, Smeeth L, Guttmann A, Harron K, Moher D, Petersen I, Sørensen HT, von Elm E, Langan SM, the RECORD Working Committee. The REporting of studies Conducted using Observational Routinely-collected health Data (RECORD) Statement. *PLoS Medicine* 2015; in press.

*Checklist is protected under Creative Commons Attribution ([CC BY](http://creativecommons.org/licenses/by/4.0/)) license.

### SM 3. Recorded clinical characteristics

The patients’ baseline clinical characteristics were extracted from the ELCAPA case report form. The variables included:

- Sociodemographic data: age, sex, educational level, living alone, requiring home assistance, place of residence (own home, family/residential care, or nursing care), number of living children, and the number of children able to provide support).
- Oncological parameters: cancer site and metastatic status.
- Frailty screening parameters: the G8 score (range: 0-17; impaired when ≤14/17), Eastern Cooperative Oncology Group Performance Status (ECOG-PS, from 0 = normal to 4 = completely disabled), asthenia, mobility, autonomy (a history of falls, time in the timed get-up-and-go test (TGUG; impaired when >20 seconds) and the activities of daily living score (ADL; range: 0-6; impaired when ≤5)), nutrition (body mass index, recent weight loss (≥10% in the last 6 months or ≥5% in the last month), cognition (Mini Mental State Examination (MMSE) score; range: 0-30; impaired when ≤23/30), mood (mini-Geriatric Depression Scale (GDS) score; range: 0-4; impaired when ≥1), comorbidities (arterial hypertension, diabetes, kidney failure, heart failure, arrythmia, depression and the Cumulative Illness Rating Scale-Geriatric (CIRS-G) score (impaired when ≥1 grade 3/4 items), and polypharmacy (number of drugs usually taken).
- The planned treatment: curative, palliative, supportive care, or no care, along with any scheduled surgical treatments, chemotherapy, and/or radiotherapy.

### SM 4. Details on the statistical analyses

***Patient dissimilarity.*** A dissimilarity metric derived from optimal matching was built using expert knowledge of the specific aspects of cancer treatment in hospital. Optimal matching is a sequence analysis method that quantifies the difference between two qualitative sequences. It was first introduced into the social sciences by Andrew Abbott (34) and has since been extended to healthcare trajectories (35-37). The analytical process consists in minimizing the sum of costs associated with unitary operations (insertion, deletion and substitution) performed to transform the first compared sequence into the other compared sequence. In our study, unitary costs were set manually by leveraging expert knowledge of specific aspects of care for older patients with cancer. Substitution costs associated with hospital admissions for cancer (especially with surgery) and cancer outpatient care were set high, in order to increase the metric’s sensitivity to variables that constitute the backbone of a cancer treatment plan. The associated insertion and deletion (indel) costs were set at the minimum possible value (half of the maximum substitution cost), in order to lower the metric’s sensitivity to the time between care episodes(38); we were more interested in gathering together patients having received the same type of treatment rather knowing the times at which they occurred. Likewise, costs associated with consultations were lowered because they are less important for hospital care trajectories. Admissions to rehabilitation units were also attributed a lesser value, in order to account for the poor representativeness of rehabilitation stays in public hospitals: a substantial proportion of rehabilitation stays take place in private clinics (39). For reproducibility purposes, details of the costs implemented per care dimension are given below.

Distances between pairs of trajectories were computed by applying the dissimilarity metric dimension-wise and aggregating with the Euclidian norm.

***Clustering.*** In order to build homogeneous clusters of care trajectories (the present study’s primary endpoint), we performed hierarchical clustering using Ward’s criterion(40) on the dissimilarity matrix. We used the Hubert’s c-index(41) and the elbow rule to determine the number of clusters with a relevant clinical meaning.

***Description of the clusters***. Clusters were subsequently described by their type, hospital care consumption intensity, and patient profiles. To do so, we computed the total number of episodes per patient-month (ppm) of follow-up within each cluster, along with the numbers of planned and unplanned (based on the admission mode) episodes ppm. The hospital length of stay (LOS) overall and for planned and unplanned admissions was computed for each cluster by summing the number of days spent in hospital for inpatient and outpatient care (consultations and emergency room visits were therefore excluded) ppm of follow-up. Care consumption indicators were compared between clusters using Wald tests on univariate Poisson analysis. Clinical features were assessed overall and pairwise between clusters by using chi-squared and Kruskal-Wallis tests for qualitative and quantitative variables, respectively. Missing data were imputed using random forests trained on the observed values; the residual mean square error and the out-of-bag error were computed as a guide to imputation performance.

***Cluster’s determinants***. Lastly, each cluster’s clinical determinants were identified in multiple one-vs-rest logistic regressions (with binary outcomes, *e.g* cluster A vs clusters B,C, and D; cluster B vs clusters A,C, and D; etc), fitted to the imputed dataset. We decided not to use multinomial logistic regression, as it requires a “reference” cluster against which the other clusters will be compared, which is not applicable to our study. Log-linearity assumptions for continuous variables were tested, and continuous predictors were modeled as categorical otherwise using the thresholds described in SM3. Potential interactions between cancer site and metastatic status were evaluated, as we expect a stratification of cancer treatment decision-making depending both on the cancer site and metastatic status. For other predictors, manual bidirectional stepwise feature selection was used. The threshold for statistical significance was set to p<0.05 and adjusted for multiple comparisons, when appropriate.

***OM costs per care dimension.***

Tables are transposable and provide the substitution cost between all pairs of subdimensions for each dimension.

1. Consultations with an HCP, indel = 1

|  | **1.0 No use** | **1.1 Other** | **1.2 Support** | **1.3 Medical** | **1.4 Oncological** |
| --- | --- | --- | --- | --- | --- |
| **1.0 No use** | 0 | 1 | 1 | 1 | 1 |
| **1.1 Other** |  | 0 | 1 | 1 | 1 |
| **1.2 Support** |  |  | 0 | 1 | 1 |
| **1.3 Medical** |  |  |  | 0 | 1 |
| **1.4 Oncological** |  |  |  |  | 0 |

2. Day hospital with a cancer indication, indel = 2

|  | **2.0 No use** | **2.1 Other** | **2.2 CT/RT** | **2.3 Surgery** |
| --- | --- | --- | --- | --- |
| **2.0 No use** | 0 | 3 | 4 | 3 |
| **2.1 Other** |  | 0 | 4 | 3 |
| **2.2 CT/RT** |  |  | 0 | 4 |
| **2.3 Surgery** |  |  |  | 0 |

3. Day hospital with a non-cancer indication, indel = 1

|  | **3.0 No use** | **3.1 One ep/month** | **3.2 Two ep/month** | **3.3 ≥3 ep/month** |
| --- | --- | --- | --- | --- |
| **3.0 No use** | 0 | 2 | 2 | 2 |
| **3.1 One ep/month** |  | 0 | 2 | 2 |
| **3.2 Two ep/month** |  |  | 0 | 2 |
| **3.3 ≥3 ep/month** |  |  |  | 2 |

4. Inpatient hospital admission with a cancer indication, indel = 3

|  | **3.0 No use** | **3.1 No surgery (home)** | **3.2 No surgery (trsf)** | **3.3 No surgery (ER)** | **3.4 Surgery (home)** | **3.5 Surgery (trsf)** | **3.6 Surgery (ER)** |
| --- | --- | --- | --- | --- | --- | --- | --- |
| **3.0 No use** | 0 | 3 | 3 | 3 | 6 | 6 | 6 |
| **3.1 No surgery (home)** |  | 0 | 3 | 3 | 6 | 6 | 6 |
| **3.2 No surgery (trsf)** |  |  | 0 | 3 | 6 | 6 | 6 |
| **3.3 No surgery (ER)** |  |  |  | 0 | 6 | 6 | 6 |
| **3.4 Surgery (home)** |  |  |  |  | 0 | 3 | 3 |
| **3.5 Surgery (trsf)** |  |  |  |  |  | 0 | 3 |
| **3.6 Surgery (ER)** |  |  |  |  |  |  | 0 |

5. Inpatient hospital admission with a non-cancer indication, indel = 1

|  | **4.0 No use** | **4.1 Home admission** | **4.2 Trsf admission** | **4.3 ER admission** |
| --- | --- | --- | --- | --- |
| **4.0 No use** | 0 | 2 | 2 | 2 |
| **4.1 Home admission** |  | 0 | 2 | 2 |
| **4.2 Trsf admission** |  |  | 0 | 2 |
| **4.3 ER admission** |  |  |  | 2 |

6. Stay in rehabilitation unit, indel = 1

|  | **6.1 No use** | **6.1 ≤14d/month** | **6.2 ≥15d/month** |
| --- | --- | --- | --- |
| **6.1 No use** | 0 | 1 | 1 |
| **6.1 ≤14d/month** |  | 0 | 1 |
| **6.2 ≥15d/month** |  |  | 0 |

7. Emergency Room visit, indel = 1

|  | **7.0 No use** | **7.1 One ep/month** | **7.2 Two ep/month** | **7.3 ≥3 ep/month** |
| --- | --- | --- | --- | --- |
| **7.0 No use** | 0 | 2 | 2 | 2 |
| **7.1 One ep/month** |  | 0 | 2 | 2 |
| **7.2 Two ep/month** |  |  | 0 | 2 |
| **7.3 ≥3 ep/month** |  |  |  | 2 |

Abbreviations: HCP, healthcare professional; CT/RT, chemotherapy and/or radiotherapy; ep/month, episode(s) per month; trsf, transfer; ER, emergency room; d/month, days per month.

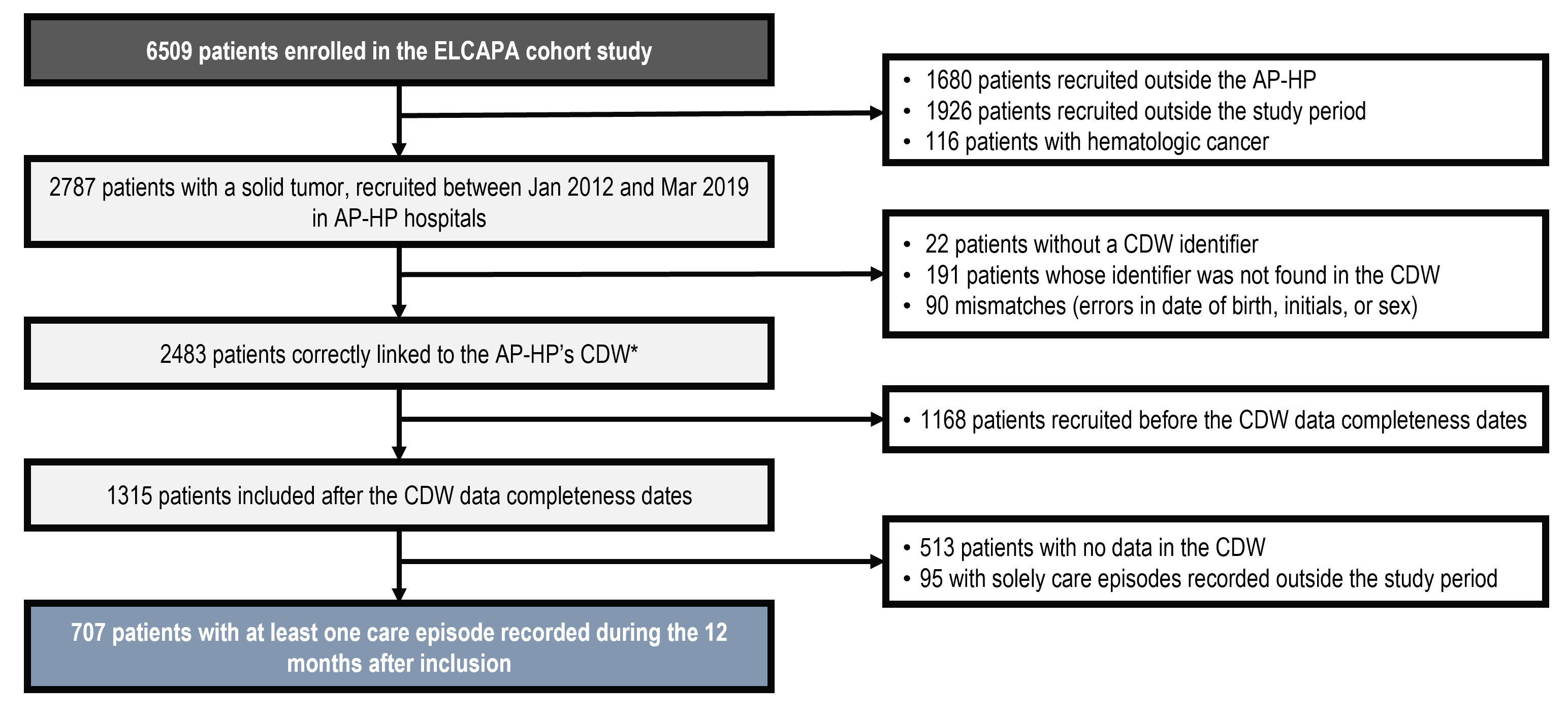

### SM5. Study flowchart

Abbreviations: ELCAPA, Elderly Cancer Patient; AP-HP, Assistance Publique-Hôpitaux de Paris (Greater Paris University Hospitals Group, Paris, France); CDW, clinical data warehouse.

*A patient was included twice in ELCAPA, and consequently had two ELCAPA’s identifiers. Deduplication was performed after the linkage with the APHP’s CDW.

### SM 6. Descriptive statistics of the included vs non-included patients

|  |  | | **Study sample** | **Study population*** | **p-value**** |
| --- | --- | --- | --- | --- | --- |
|  |  | | N=707 | N=2080 |  |
| **Age in years, median (IQR), missing(M)=2** | | | 82.0 [78.0;86.0] | 82.0 [78.0;86.0] | 0.052 |
| **Sex (male), N (%)** | | | 370 (52.3) | 1075 (51.7) | 0.798 |
| **Metastatic cancer, N (%), M=239** | | | 311 (45.2) | 940 (50.5) | **0.019** |
| **Cancer site, N (%), M=14** | | Breast/prostate | 168 (23.9) | 518 (25.0) | 0.500 |
|  |  | Colorectal | 128 (18.2) | 345 (16.7) |  |
|  |  | Other digestive tract site | 134 (19.1) | 366 (17.7) |  |
|  |  | Urinary tract | 95 (13.5) | 309 (14.9) |  |
|  |  | Lung/ENT | 63 (8.97) | 222 (10.7) |  |
|  |  | Other | 114 (16.2) | 311 (15.0) |  |
| **ECOG-PS, N (%), M=22** | | 0/1 | 322 (45.9) | 1018 (49.3) | 0.291 |
|  |  | 2 | 175 (25.0) | 491 (23.8) |  |
|  |  | 3/4 | 204 (29.1) | 555 (26.9) |  |
| **Asthenia, N (%), M=35** | | | 489 (69.8) | 1496 (72.9) | 0.116 |
| **Impaired G8 (<=14), N (%), M=471** | | | 579 (86.3) | 1426 (86.7) | 0.851 |
| **History of falls, N (%), M=59** | | | 193 (27.8) | 590 (29.0) | 0.580 |
| **Impaired ADL (<=5), N (%), M=43** | | | 185 (26.4) | 594 (29.1) | 0.181 |
| **TGUG <= 20s, N (%), M=783** | | | 362 (68.0) | 1025 (69.6) | 0.532 |
| **Impaired MMSE (<=23), N (%), M=496** | | | 154 (24.5) | 490 (29.5) | **0.020** |
| **Weight loss , N (%)** (≥10% in last 6 months or ≥5% in last month) | | | 98 (13.9) | 353 (17.0) | 0.060 |
| **Impaired Mini-GDS (>=1), N (%), M=376** | | | 224 (33.4) | 608 (34.9) | 0.521 |
| **Depression (comorbidity), N (%), M=1894** | | | 110 (30.6) | 147 (27.6) | 0.374 |
| **Impaired CIRS-G (>=1 grade 3/4), N (%), M=210** | | | 469 (70.0) | 1335 (70.0) | 1.000 |
| **Number of drugs, median (IQR), M=128** | | | 7.00 [4.00;9.00] | 6.00 [4.00;8.00] | **<0.001** |
| **Comorbidities, N (%)** | | HT, M=19 | 487 (69.3) | 1391 (67.4) | 0.373 |
|  |  | Diabetes, M=28 | 183 (26.1) | 459 (22.3) | **0.045** |
|  |  | Kidney failure, M=166 | 275 (43.0) | 780 (39.4) | 0.109 |
|  |  | Heart failure, M=87 | 86 (12.9) | 218 (10.7) | 0.151 |
|  |  | Arrythmia, M=94 | 180 (26.7) | 535 (26.5) | 0.977 |
| **Living alone, N (%), M=14** | | | 257 (36.5) | 824 (39.8) | 0.130 |
| **≥1 available children, N (%), M=587** | | | 475 (83.9) | 1382 (84.6) | 0.762 |
| **Place of residence, N (%), M=10** | | Family/residential home | 49 (6.96) | 131 (6.32) | 0.662 |
|  |  | Nursing care or other | 29 (4.12) | 99 (4.78) |  |
|  |  | Own home | 626 (88.9) | 1843 (88.9) |  |
| **Home assistance, N (%), M=45** | | | 327 (47.3) | 983 (47.9) | 0.817 |
| **Planned treatment, N(%), M=596** | | Curative care | 332 (51.9) | 690 (44.5) | **<0.001** |
|  |  | Palliative care | 241 (37.7) | 572 (36.9) |  |
|  |  | Supportive care/No care | 67 (10.5) | 289 (18.6) |  |
| Abbreviations: IQR, interquartile range; ENT, ear nose throat; ECOG-PS, performance status; ADL, activities of daily living; TGUG, timed get-up and go; MMSE, Mini Mental State Examination; GDS, Geriatric Depression Scale; CIRS-G, Cumulative Illness Rating Scale - Geriatric; HT, hypertension; CT/RT, chemotherapy/radiotherapy *Patients with a solid tumor, recruited between Jan 2012 and Mar 2019 in AP-HP hospitals and meeting at least one subsequent exclusion criteria. **Kruskal-Wallis and Chi-squared tests for quantitative and qualitative features respectively, global comparison. | | | | | |

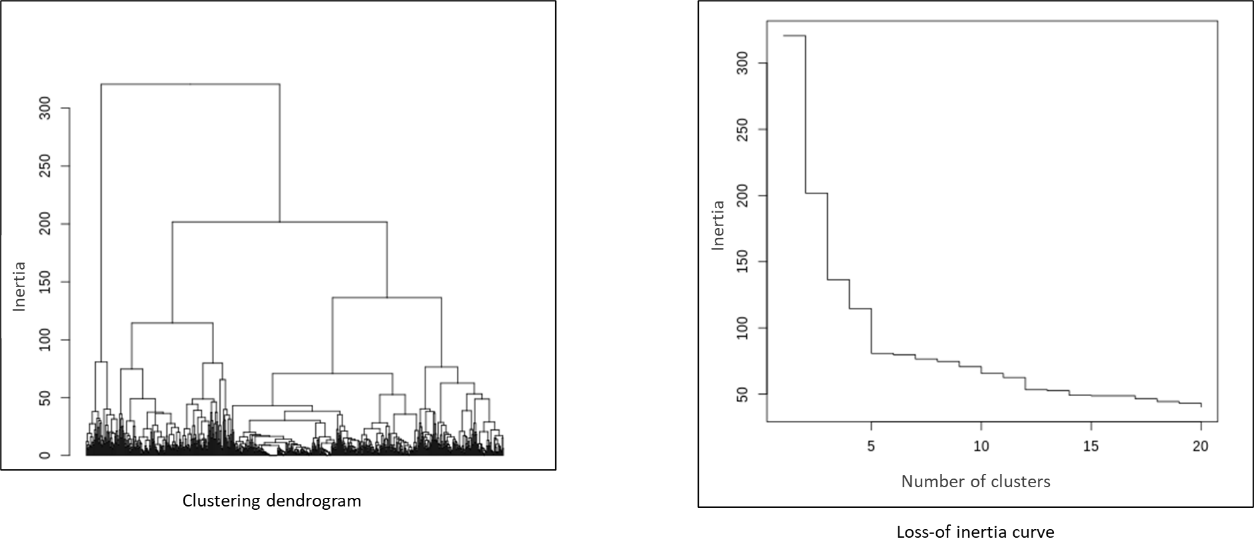

### SM 7. The clustering dendrogram and loss-of-inertia curve

### SM 8. Sensitivity analysis on newly diagnosed cancer patients only

We restricted our study sample to newly diagnosed patients at the time of geriatric assessment (n=503). We performed the same methodology, which resulted in similar clusters as the principal analysis : Cluster A (n=129, 25.6%) with in-hospital surgical trajectories, Cluster B (n=113, 22.5%) with outpatient trajectories made of mixed chemotherapy and radiotherapy, Cluster C (n=217, 43.1%) with no hospital cancer treatment, and Cluster D (n=44, 8.7%), with chemotherapy-dominant trajectories and high consumption of hospital care (figure not shown). Corresponding clinical profiles are given below.

| **SM10. Clinical features and care consumption, by cluster** | | | | |  |  |
| --- | --- | --- | --- | --- | --- | --- |
|  |  | **Cluster A** | **Cluster B** | **Cluster C** | **Cluster D** | **p-value*** |
|  |  | **N=129** | **N=113** | **N=217** | **N=44** |  |
| **Age in years, median (IQR)** | | 82.0 [78.0;85.0] | 82.0 [79.0;86.0] | 84.0 [80.0;88.0] | 80.0 [73.8;85.0] | 0.001 |
| **Sex (male), N (%)** | | 63 (48.8%) | 52 (46.0%) | 112 (51.6%) | 23 (52.3%) | 0.781 |
| **Metastatic cancer, N (%), M=13** | | 24 (18.9%) | 55 (49.5%) | 99 (47.6%) | 21 (47.7%) | <0.001 |
| **Cancer site, N (%), M=2** | Breast/prostate | 23 (18.0%) | 29 (25.7%) | 41 (19.0%) | 10 (22.7%) |  |
|  | Colorectal | 42 (32.8%) | 16 (14.2%) | 34 (15.7%) | 8 (18.2%) |  |
|  | Other digestive tract site | 29 (22.7%) | 18 (15.9%) | 57 (26.4%) | 16 (36.4%) |  |
|  | Urinary tract | 18 (14.1%) | 12 (10.6%) | 23 (10.6%) | 2 (4.55%) |  |
|  | Lung/ENT | 6 (4.69%) | 17 (15.0%) | 24 (11.1%) | 2 (4.55%) |  |
|  | Other | 10 (7.81%) | 21 (18.6%) | 37 (17.1%) | 6 (13.6%) |  |
| **ECOG-PS, N (%), M=2** | 0/1 | 80 (62.0%) | 54 (48.2%) | 65 (30.1%) | 29 (65.9%) | <0.001 |
|  | 2 | 16 (12.4%) | 32 (28.6%) | 59 (27.3%) | 8 (18.2%) |  |
|  | 3/4 | 33 (25.6%) | 26 (23.2%) | 92 (42.6%) | 7 (15.9%) |  |
| **Asthenia, N (%), M=2** | | 77 (60.2%) | 78 (69.6%) | 165 (76.0%) | 28 (63.6%) | 0.016 |
| **Impaired G8 (<=14), N (%), M=23** | | 103 (82.4%) | 91 (82.7%) | 191 (94.1%) | 32 (76.2%) | 0.001 |
| **History of falls, N (%), M=8** | | 32 (25.0%) | 20 (17.9%) | 81 (38.4%) | 12 (27.3%) | 0.001 |
| **Impaired ADL (<=5), N (%), M=2** | | 24 (18.6%) | 23 (20.4%) | 82 (38.1%) | 8 (18.2%) | <0.001 |
| **TGUG <= 20s, N (%), M=120** | | 75 (77.3%) | 64 (70.3%) | 84 (53.5%) | 29 (76.3%) | <0.001 |
| **Impaired MMSE (<=23), N (%), M=60** | | 29 (24.6%) | 20 (20.0%) | 58 (31.2%) | 11 (28.2%) | 0.210 |
| **Weight loss , N (%)** (≥10% in last 6 months or ≥5% in last month) | | 9 (6.98%) | 14 (12.4%) | 41 (18.9%) | 6 (13.6%) | 0.019 |
| **Impaired Mini-GDS (>=1), N (%), M=27** | | 37 (29.8%) | 30 (28.0%) | 90 (44.1%) | 8 (19.5%) | 0.001 |
| **Depression (comorbidity), N (%), M=252** | | 21 (32.8%) | 12 (19.7%) | 44 (40.7%) | 6 (33.3%) | 0.050 |
| **Impaired CIRS-G (>=1 grade 3/4), N (%), M=23** | | 76 (61.3%) | 74 (69.2%) | 162 (77.9%) | 22 (53.7%) | 0.001 |
| **Number of drugs, median (IQR), M=19** | | 7.00 [4.00;9.00] | 7.00 [4.00;9.00] | 7.00 [5.00;9.00] | 6.00 [3.00;9.00] | 0.562 |
| **Comorbidities, N (%)** | HT, M=2 | 96 (74.4%) | 75 (66.4%) | 149 (69.3%) | 32 (72.7%) | 0.550 |
|  | Diabetes, M=2 | 43 (33.6%) | 28 (24.8%) | 58 (26.9%) | 11 (25.0%) | 0.406 |
|  | Kidney failure, M=51 | 42 (35.6%) | 52 (52.0%) | 96 (50.0%) | 17 (40.5%) | 0.039 |
|  | Heart failure, M=25 | 20 (15.9%) | 13 (12.0%) | 32 (16.0%) | 4 (9.09%) | 0.546 |
|  | Arrythmia, M=20 | 34 (27.6%) | 25 (22.5%) | 58 (28.3%) | 9 (20.5%) | 0.546 |
| **Living alone, N (%)** | | 47 (36.4%) | 44 (38.9%) | 86 (39.6%) | 14 (31.8%) | 0.769 |
| **≥1 available children, N (%), M=101** | | 86 (86.0%) | 84 (84.8%) | 128 (79.5%) | 38 (92.7%) | 0.171 |
| **Place of residence, N (%)** | Family/residential home | 9 (6.98%) | 8 (7.08%) | 14 (6.45%) | 5 (11.4%) | 0.363 |
|  | Nursing care or other | 2 (1.55%) | 6 (5.31%) | 14 (6.45%) | 1 (2.27%) |  |
|  | Own home | 118 (91.5%) | 99 (87.6%) | 189 (87.1%) | 38 (86.4%) |  |
| **Home assistance, N (%), M=12** | | 49 (38.3%) | 58 (52.7%) | 111 (53.1%) | 16 (36.4%) | 0.016 |
| Abbreviations: IQR, interquartile range; ENT, ear nose throat; ECOG-PS, performance status; ADL, activities of daily living; TGUG, timed get-up and go; MMSE, Mini Mental State Examination; GDS, Geriatric Depression Scale; CIRS-G, Cumulative Illness Rating Scale - Geriatric; HT, hypertension *Kruskal-Wallis and Chi-squared tests for quantitative and qualitative features respectively, global comparison. | | | | | | |

### SM 9. Further description of cluster C (“Recurrent unplanned care without any hospital cancer treatment”)

Cluster C’s chronogram could also be split visually into two subgroups by death status, even if this separation was not done automatically during the clustering process (a separation in five instead of four clusters would have further cut cluster B in two subgroups) . Indeed, the 168 (55.6% of cluster C) patients who died during follow-up (Table below) consumed high levels of inpatient care (especially unplanned stays) before they died, while the 134 (44.4% of cluster C) patients who were still alive at the end of the study had a very low level of hospital care consumption (mostly regular consultations). With regard to clinical features, we found that the patients who were still alive at the end of the study had higher proportions of breast or prostate cancer (35.3% vs. 15.7% among patients who died during follow-up) and a lower proportion of metastatic cancer (38.2% vs. 64.6% respectively). The patients who were still alive also had better geriatric parameters, in general.

|  |  | **Alive** | **Dead** | **p-value*** |
| --- | --- | --- | --- | --- |
|  |  | N=134 | N=168 |  |
| **Age in years, median (IQR)** | | 82.0 [77.2;85.8] | 83.0 [80.0;88.0] | **0.022** |
| **Sex (male), N (%)** | | 58 (43.3) | 99 (58.9) | **0.010** |
| **Metastatic cancer, N (%), missing(M)=10** | | 50 (38.2) | 104 (64.6) | **<0.001** |
| **Cancer site, N (%), M=3** | Breast/prostate | 47 (35.3) | 26 (15.7) | **0.003** |
|  | Colorectal | 19 (14.3) | 23 (13.9) |  |
|  | Other digestive tract site | 26 (19.5) | 39 (23.5) |  |
|  | Urinary tract | 13 (9.77) | 24 (14.5) |  |
|  | Lung/ENT | 8 (6.02) | 20 (12.0) |  |
|  | Other | 20 (15.0) | 34 (20.5) |  |
| **ECOG-PS, N (%), M=4** | 0/1 | 70 (52.6) | 35 (21.2) | **<0.001** |
|  | 2 | 32 (24.1) | 51 (30.9) |  |
|  | 3/4 | 31 (23.3) | 79 (47.9) |  |
| **Asthenia, N (%), M=3** | | 88 (66.2) | 140 (84.3) | **<0.001** |
| **Impaired G8 (<=14), N (%), M=27** | | 107 (83.6) | 144 (98.0) | **<0.001** |
| **History of falls, N (%), M=9** | | 36 (27.3) | 65 (40.4) | **0.026** |
| **Impaired ADL (<=5), N (%), M=4** | | 31 (23.3) | 71 (43.0) | **0.001** |
| **TGUG <= 20s, N (%), M=86** | | 63 (61.2) | 63 (55.8) | 0.504 |
| **Impaired MMSE (<=23), N (%), M=39** | | 27 (23.1) | 51 (34.9) | 0.050 |
| **Weight loss , N (%)** (≥10% in last 6 months or ≥5% in last month) | | 11 (8.21) | 40 (23.8) | **0.001** |
| **Impaired Mini-GDS (>=1), N (%), M=17** | | 34 (26.4) | 85 (54.5) | **<0.001** |
| **Depression (comorbidity), N (%), M=141** | | 23 (28.7) | 35 (43.2) | 0.081 |
| **Impaired CIRS-G (>=1 grade 3/4), N (%), M=15** | | 92 (72.4) | 129 (80.6) | 0.135 |
| **Number of drugs, median (IQR), M=12** | | 6.00 [4.00;8.25] | 7.00 [5.00;9.00] | 0.054 |
| **Comorbidities, N (%)** | AHT, M=4 | 97 (73.5) | 111 (66.9) | 0.267 |
|  | Diabetes, M=4 | 32 (24.2) | 43 (25.9) | 0.846 |
|  | Kidney failure, M=36 | 50 (44.2) | 74 (48.4) | 0.588 |
|  | Heart failure, M=23 | 15 (12.0) | 24 (15.6) | 0.493 |
|  | Arrythmia, M=17 | 30 (23.4) | 51 (32.5) | 0.121 |
| **Living alone, N (%), M=2** | | 54 (40.6) | 62 (37.1) | 0.621 |
| **≥1 available children, N (%), M=72** | | 87 (87.0) | 102 (78.5) | 0.133 |
| **Place of residence, N (%), M=2** | Family/residential home | 11 (8.27) | 7 (4.19) | 0.229 |
|  | Nursing care or other | 6 (4.51) | 12 (7.19) |  |
|  | Own home | 116 (87.2) | 148 (88.6) |  |
| **Home assistance, N (%), M=10** | | 58 (45.0) | 91 (55.8) | 0.084 |
| **Follow-up time in days, median (IQR)** | | 258 [72.5;322] | 103 [51.8;199] | **<0.001** |
| **Number of care episodes (ppm), median (IQR)** | Total | 1.06 [1;1.12] | 1.46 [1.38;1.54] |  |
|  | Planned | 0.99 [0.93;1.05] | 1.27 [1.2;1.35] |  |
|  | Unplanned | 0.07 [0.06;0.09] | 0.19 [0.16;0.22] |  |
| **Hospital LOS (ppm), median (IQR)** | Total | 2.52 [2.43;2.62] | 7.88 [7.7;8.07] |  |
|  | Planned | 2.09 [2;2.18] | 5.51 [5.36;5.67] |  |
|  | Unplanned | 0.43 [0.39;0.48] | 2.37 [2.27;2.47] |  |
| **Consultations** | N patients (%) | 134 (100) | 158 (94.0) | **0.003** |
|  | N episodes ppy | 9.76 [9.11;10.45] | 10.2 [9.48;10.96] |  |
| **DH - Cancer** | N patients (%) | 34 (25.4) | 49 (29.2) | 0.546 |
|  | N episodes ppy | 0.59 [0.44;0.78] | 1.51 [1.24;1.82] |  |
|  | CT - N patients (%) | 7 (5.22) | 17 (10.1) | 0.177 |
|  | CT - N episodes ppy | 2.68 [1.43;4.58] | 6.32 [4.59;8.48] |  |
|  | RT - N patients (%) | 5 (3.73) | 22 (13.1) | **0.009** |
|  | RT - N episodes ppy | 3.78 [2.16;6.13] | 6.25 [4.65;8.21] |  |
| **DH - No Cancer** | N patients (%) | 15 (11.2) | 17 (10.1) | 0.910 |
| **INPT - Cancer** | N patients (%) | 50 (37.3) | 131 (78.0) | **<0.001** |
|  | N patients (%) - planned | 43 (32.1) | 99 (58.9) | **<0.001** |
|  | N patients (%) - unplanned | 15 (11.2) | 66 (39.3) | **<0.001** |
|  | N patients (%) - surgery | 13 (9.70) | 9 (5.36) | 0.222 |
| **INPT - No Cancer** | N patients (%) | 45 (33.6) | 60 (35.7) | 0.791 |
|  | N patients (%) - planned | 30 (22.4) | 39 (23.2) | 0.974 |
|  | N patients (%) - unplanned | 19 (14.2) | 29 (17.3) | 0.569 |
| **Rehabilitation** | N patients (%) | 17 (12.7) | 36 (21.4) | 0.067 |
| **ER** | N patients (%) | 27 (20.1) | 32 (19.0) | 0.925 |
|  | N episodes ppy | 0.42 [0.29;0.58] | 0.57 [0.41;0.77] |  |
| Abbreviations: IQR, interquartile range; ENT, ear nose throat; ECOG-PS, performance status; ADL, activities of daily living; TGUG, timed get-up and go; MMSE, Mini Mental State Examination; GDS, Geriatric Depression Scale; CIRS-G, Cumulative Illness Rating Scale - Geriatric; AHT, hypertension; LOS, length of stay; ppy, per patient-year ; DH, day hospital; CT, chemotherapy; RT, radiotherapy; INPT, inpatient; ER, emergency Room *Kruskal-Wallis, Wald and Chi-squared tests for quantitative, episode counts and qualitative features respectively, global comparison. | | | | |
